# The accuracy of saliva versus nasopharyngeal and/or oropharyngeal samples for the detection of SARS-CoV-2 in children – A rapid systematic review and meta-analysis

**DOI:** 10.1101/2021.06.21.21259284

**Authors:** Sharonjit K. Dhillon, Petra Schelstraete, Laura Cornelissen, Yves Lafort, Jesper Bonde, M. Arbyn

**Affiliations:** Department of Epidemiology and Public Health, Sciensano, Brussels, Belgium; UZ Gent, Department of Paediatric Pulmonology and Infectious Diseases; Molecular Pathology Laboratory, Dept. Pathology, Copenhagen University Hospital - Amager and Hvidovre Hospital, Copenhagen, Denmark

**Keywords:** SARS-CoV-2, children, diagnostic test accuracy, saliva, COVID-19

## Abstract

**Background:** The comparative performance of saliva and nasopharyngeal samples for the detection of severe acute respiratory syndrome coronavirus 2 (SARS-CoV-2) infection by reverse transcriptase polymerase chain reaction (RT-PCR) in children remains unclear. As schools reopen around the world, there is an interest in the use of saliva samples for the detection of SARS-CoV-2 in children to circumvent barriers with nasopharyngeal sampling. We systematically reviewed the literature to understand the performance of saliva sampling using RT-PCR on naso- and/or oropharyngeal swabs as the reference standard.

**Methods:** Articles from PubMed/MEDLINE and Living Evidence were accessed until 28^th^ April 2021. A search method without restriction to children population was applied and during the review phase, if a study included patients <18 years old, authors were contacted to provide additional information on the subset of children. Studies were eligible if they reported on matched saliva and naso- and/or oropharyngeal samples, taken from the same patient on the same day. Studies using other respiratory samples such as sputum samples were excluded. Each paired patient sample had to be tested on the same RT-PCR platform.

**Results:** Ten studies were included, comprising 1486 matched saliva and on naso- and/or oropharyngeal pairs from children aged 0 to 18 years old. The pooled absolute sensitivity and specificity of saliva sampling using RT-PCR on nasopharyngeal samples as the reference standard was 84.5% (95% CI; 78.0%-90.3%) and 99.5% (95% CI; 98.2%-100.0%). Comparable performance of saliva to nasopharyngeal samples was shown in both symptomatic and asymptomatic children. Stratified analyses of various covariates showed no significant differences.

**Discussion:** Our pooled accuracy estimates of RT-PCR SARS-CoV-2 testing on saliva in children did not seem to be different from meta-analyses of studies that enrolled mainly adults. Saliva could potentially be considered an alternative sampling method for screening in children and to pick up those with high viral load.

## Introduction

The severe acute respiratory syndrome coronavirus 2 (SARS-CoV-2) was initially reported in Wuhan, China at the end of 2019. Since then, the world has witnessed the devastating impact of this pandemic with over 2.75 million deaths and 123 million cases as of March 2021^1^. In the midst of this pandemic, children and adolescents are seen as a distinct population as they rarely develop severe or critical illness. Compared to adults, children infected with SARS-CoV-2 are less likely to exhibit symptoms or have mild, non-specific symptoms^2,3^. Additionally, despite a majority of children exhibiting mild disease, children can actively transmit SARS-CoV-2 to others. With the reopening of schools worldwide, there is an interest in testing in children for SARS-CoV-2 to identify new clusters of infection early and prevent outbreaks and unnecessary school closures. Hence, the need for easy and repeated sampling is essential to ensure this.

Nasopharyngeal swabs (NPS) for nucleic acid amplification testing (NAAT) are regarded as the gold standard for the identification of SARS-CoV-2. NPS are obtained through the insertion of swabs with the purpose to reach the deepest area of the nasopharynx. This invasive method sometimes is difficult and uncomfortable, especially in children. In general, children are often uncooperative and may be anxious during sampling, increasing the risk of trauma and unwillingness to be tested. On top of that, NPS sampling requires trained healthcare personnel for collection and can increase the risk of nosocomial viral transmission as the procedure often induces cough and sneezing.

Alternate sampling methods, specifically saliva, have been the subject of recent research. The ease and non-invasive nature of saliva collection and the possibility of self-sampling have the potential to address many of the barriers associated with NPS sampling, especially in paediatric populations. Hence, there is a need to understand the comparative sensitivity of saliva and the NPS for SARS-CoV-2 detection in children. A preliminary search on public bibliographic databases, including peer-reviewed and preprint articles showed that paediatric evidence for the use of saliva specimens by RT-PCR is sparse and suffers from methodological limitations. Therefore, we contacted authors of eligible studies involving children and requested outcome data for paediatric strata. These data allowed us to conduct for the first time a systematic review and meta-analysis on the accuracy of SARS-CoV-2 assays applied on paired saliva and NPS in children.

## Methods

### Search strategy and selection criteria

A search method without restriction to population was applied. During the review phase, if a study included patients <18 years old, authors were contacted to provide additional information on the subset of the children population. We searched the databases PubMed/MEDLINE and Living Evidence (see appendix for the clinical questions and search terms used). Living Evidence is an initiative from the University of Bern which includes live search results using application programming interfaces (API) to collect daily citation data from peer-reviewed medical bibliographic databases (PUBMED, Embase) as well as preprints indexed in *bioRxiv* and *medRxiv* databases. The applied search strings are in the appendix (p 1). Additionally, relevant studies were retrieved by screening the reference lists of searched studies. Studies were eligible for inclusion if the following criteria were fulfilled: a matched saliva and naso- and/or oropharyngeal sample taken from the same patient on the same day. Studies using other respiratory, samples such as sputum samples, were excluded. Each paired patient sample had to be tested on the same RT-PCR platform. Where primary data was not included in the article, we contacted authors to provide additional accuracy data whenever possible.

For this systematic review and meta-analysis, we followed the PRISMA guidelines for reporting meta-analyses^4^. Additional details on the Population-Intervention-Comparator-Outcome-Target Condition-Study (PICOTS) and the review question are described in the appendix (p 2).

### Clinical questions, data extraction and quality assessment

We aimed to answer the following research question: what is the absolute sensitivity and specificity of saliva compared to the standard nasopharyngeal samples for the detection of SARS-CoV-2 in children? We analysed studies including symptomatic and asymptomatic children who were tested for SARS-CoV-2 infection and children undergoing screening for SARS-CoV-2 infection as part of a screening programme or as close contacts of a confirmed COVID-19 case. Children with confirmed COVID-19 undergoing retesting were also included. Two authors (SKD and MA) independently screened articles and extracted the data using a standardized data sheet, which was completed with information on study country, population and symptom status, age, index specimen collected (self-collected or supervised), the technique used to collect index specimen, comparator specimen, the time between sample collection and testing, device and medium used for index and comparator samples, SARS-CoV-2 assay used and gene targets. Patient-paired data was used to construct 2×2 contingency tables. Risk of bias and applicability concerns were assessed using the adapted Quality Assessment of Diagnostic Accuracy Studies 2 (QUADAS-2) tool^5^. The two reviewers assessed the findings for agreement and disagreements were solved through consensus after discussion.

### Statistical analysis

Stata (Stat version 16, StataCorp LLC, College Station, TX, USA) was used to conduct meta-analytical pooling using random effect models of binomial data. Pool proportions were analysed with the command *metaprop*^*6*^. We produced results of the systematic review in forest plots. A continuity correction was applied when a cell of the contingency table contained zero. Heterogeneity among studies was assessed by Cochran’s Q-test (p-value below 0.05 was defined as statistically significant) and by the inconsistency index (I^2^) which describes the proportion of total heterogeneity due to inter-study variation^7^. To investigate possible factors to heterogeneity, we conducted sub-group meta-analyses. We analysed the following covariates; symptom status, the method used for saliva collection and variation between pre-print and peer-reviewed studies. We considered the heterogeneity in the study findings through visual inspection of forest plots and by the p-value of inter-group heterogeneity. Statistical tests were two-sided and statistical significance was defined as p values of less than 0.05.

## Results

We identified 518 articles (published and preprints) of which 415 were excluded after review of title and abstract **(Figure 1)**. 103 papers were retained and assessed for full-text eligibility and 89 papers were excluded as they did not include children (individuals aged <18 years). Out of the 14 papers^8-21^ which fulfilled our eligibility criteria, we requested additional accuracy data for 9 papers of which we obtained 5. The final 10^8,10-17,21^ studies included in the meta-analysis comprised 5 studies for which data was received from authors and 5 studies where data was available in the original article. Two studies^12,15^ included 2 separate datasets each, hence a total number of 12 datasets from 10 studies were included in the final analyses.

**Figure 1.**
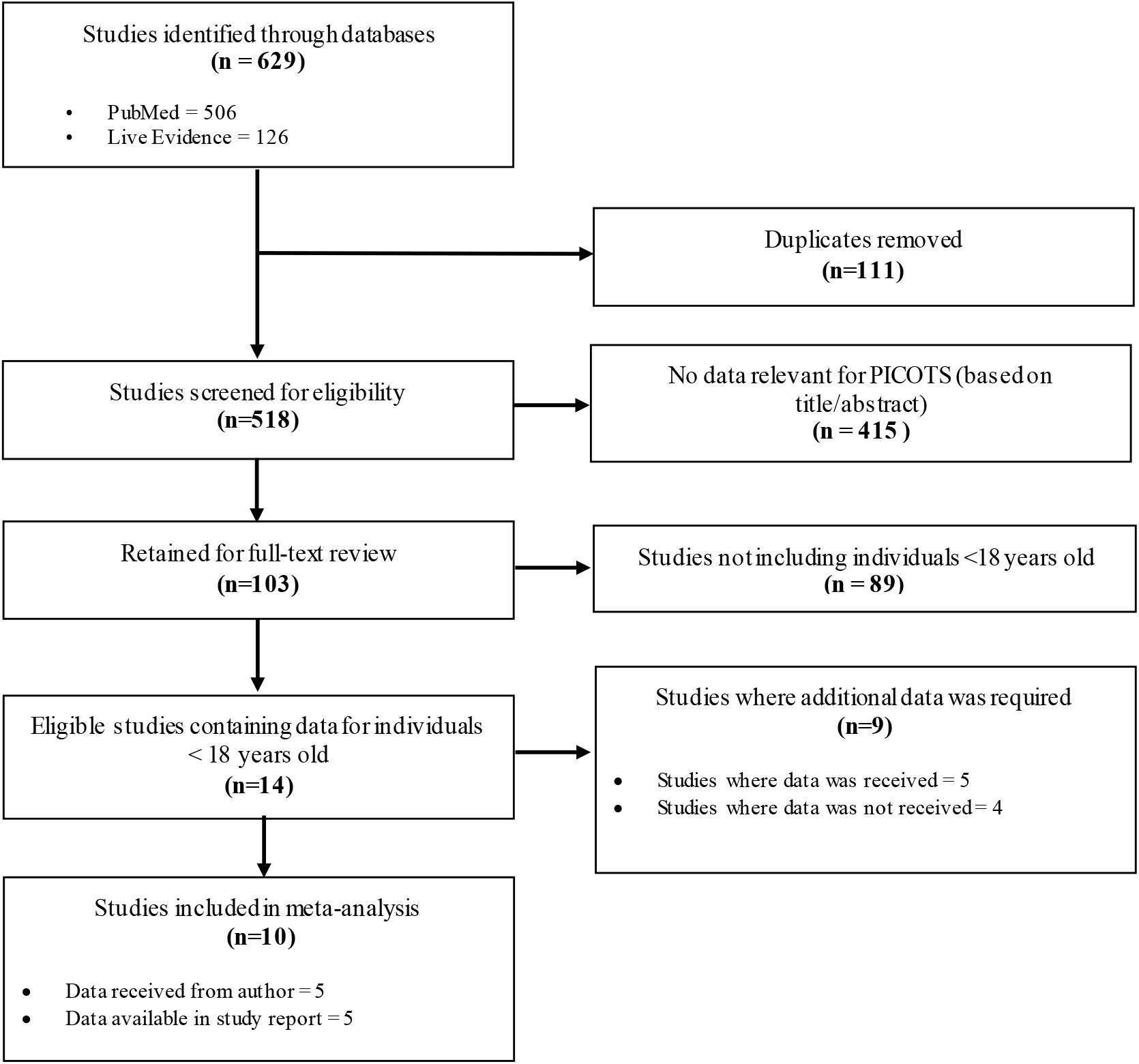
PRISMA Flow Diagram

### Description of included studies

The 10 included studies comprised of 6 peer-reviewed and 4 pre-print articles. In total, 1486 patients with matched saliva and nasopharyngeal swabs were included. The studies varied by setting and patient population, with four studies from Europe, two from Asia, two from the US and one study each from Canada and Brazil. The age of the included children ranged from 1 to 18 years.

Most studies included outpatient individuals being screened for SARS-CoV-2 infection who were either symptomatic or/and asymptomatic but were close contacts of a SARS-CoV-2 positive case. However, one study included confirmed COVID-19 hospitalised patients^16^ undergoing retesting. Three studies had only symptomatic patients and one study only asymptomatic ones. The remaining six studies included both asymptomatic and symptomatic patients, but only one study^17^ provided stratified data based on symptom status. All of the studies utilised nasopharyngeal swabs collected by healthcare workers as the comparator test except for two studies^10,14^ which included nasopharyngeal and/or oropharyngeal swabs. Different techniques for saliva collection were used: general spitting technique in 5 studies^8,11,12,15,21^, drooling method in one study^13^, posterior oropharyngeal spitting technique (by asking children to clear their throats thoroughly and collect saliva) in one study^17^ and 3 studies did not report the saliva collection technique^10,14,16^. In addition to the usual saliva collection, one study collected saliva after oropharyngeal washing^12^ (where patients were asked to gargle 2ml of saline solution 1-2 minutes prior to saliva collection). In another study^15^, gargle samples were collected after children were asked to swish 5ml of 0.9% saline solution in their mouths and the contents were emptied into a container (full technique described in **Table 1**).

**Table 1.** Study characteristic of included studies

| Study | Country | Age range | Population & symptomatic status | Number of paired samples | Saliva collection | Collection of saliva sample | Reference specimen |
| --- | --- | --- | --- | --- | --- | --- | --- |
| Felix<sup>†</sup><br>2021 | Brazil | Mean age<br>10.24 years | -Outpatients undergoing SARS-CoV-2 testing<br>-Mild symptomatic patients | n=50 | Saliva | Children were asked to spit into a sterile container for collection of about 1ml of saliva | Nasopharyngeal swabs |
| Fougère<sup>†</sup><br>2021 | Switzerland | 1-18 years<br>Mean age<br>12.7 years | -Outpatients undergoing SARS-CoV-2 testing<br>-Symptomatic patients | n=397 | Saliva | Saliva was collected by drooling method<br>Volume of saliva collected not reported | Nasopharyngeal swabs |
| Goldfarb<br>2021 | Canada | 4-14 years | -Patients presenting at outpatient centre for SARS-CoV-2 testing<br>-Symptomatic patients | n=7<br>(Saliva/NPS)<br>n=14<br>(Gargle samples/NPS) | 1. Saliva<br>(Self-collected)<br>2. Gargle Samples | 1. For saliva, children were asked to pool and spit saliva repeatedly until at least 5 – 10 ml were collected<br>2. Gargle samples were collected after contents of 5ml of 0.9% saline were squeezed into child's open mouth. Children were asked to swish contents for 5 seconds followed by tilting their heads back and gargling for 5 seconds. Process was repeated 2 more times and saline was emptied into empty container. | Nasopharyngeal swabs |
| Gonzalez<br>2021 | Spain | 4 -14 years | - Outpatients symptomatic & asymptomatic undergoing SARS-CoV-2 testing<br>- Asymptomatic 42, symptomatic 55 (data not stratified) | n=41<br>(Saliva/NPS pairs)<br>n=56<br>(Oropharyngeal washing/NPS pairs) | 1. Saliva<br>(supervised-collection)<br>2. Oropharyngeal washing | 1. Saliva specimen collected by asking patients to repeatedly spit up to a minimum of 1 ml of saliva into the collection pot<br>2. Oropharyngeal washing with 2 ml of saline solution for 1-2 minutes prior to the collection | Nasopharyngeal swabs |
| Huber<br>2021 | Switzerland | 5-17 years | -Outpatients undergoing SARS-CoV-2 testing<br>- Symptomatic & asymptomatic (data stratified) | n=170 | Saliva*<br>(Self-collected) | Individuals were asked to clear their throat thoroughly and collect saliva one or two times into the same tube (0.5-1ml)<br>Enhanced saliva collection : individuals were asked to clear their throat three times and collect saliva into the same tube (0.5-1ml) | Nasopharyngeal swabs |
| Suwaidi 2021 | UAE | 3-18 years | -Community based screening centres for closed contacts with confirmed COVID-19 patients, presence of presumptive symptoms or testing for return to school.<br>-Symptomatic & asymptomatic (data not stratified) | n=485 | Saliva (Self-collected) | Participants were asked to close their mouths and allow saliva to pool in mouth for 1-2 minutes and gently spit 1-3 ml of saliva in the container | Nasopharyngeal swabs |
| Delaney<sup>†</sup> 2020 | USA | 9-20 years | -Individuals tested for COVID-19 in emergency department and peri-operative testing program<br>-41 asymptomatic, 5 symptomatic (data not stratified) | n=46 | Saliva | 1-2 ml saliva collected. Collection technique not reported. | Nasopharyngeal & oropharyngeal swabs |
| Gavars<sup>†</sup> 2020 | Latvia | 5-17 years | -Individuals who were contact persons of confirmed COVID-19 persons.<br>-All asymptomatic | n=125 | Saliva (Self-collected) | NR | Nasopharyngeal & oropharyngeal swabs |
| Han 2020 | South Korea | <18 years | -In patient confirmed COVID-19 patients<br>-Symptomatic & asymptomatic | n=11 | Saliva | NR | Nasopharyngeal swabs |
| Yee 2020 | USA | 4-17 years | -In-patient, out-patient and household members of previously diagnosed COVID-19 positive individuals<br>-Symptomatic & asymptomatic | n=93 | Saliva (Self-collected) | 3 ml of saliva was self-collected under observation of HCW, patients were asked to work up saliva by gently rubbing the outside of their cheeks and gently spitting without coughing or clearing their throats | Nasopharyngeal swabs |

The volume of saliva collected was less than 2ml in four studies, between 2-4ml in two studies and more than 5ml of saliva in two studies. Four studies did not report the volume of saliva collected for molecular testing.

Although saliva collection is more comfortable than NPS sampling in children, obtaining an appropriate saliva specimen still can be challenging, particularly among infants. Delaney et al^10^ reported that 2% of patients were unable to provide sufficient volume (1-2ml) of saliva for testing. The authors recommended the age cut off of five years and older for saliva sampling as many of those younger than five were unable to provide saliva samples for testing. They also observe that children over the age of five years portrayed the developmental maturity to provide a sufficient volume of saliva whereas those younger than five years required more time. Difficulty in obtaining saliva samples was also reported by Fernández-González et al^12^ where 54 patients with a median age of 5.8 years old were unable to provide saliva specimens. Transport media for saliva specimens were used in two studies whereas six studies collected saliva specimens without transport medium. Nine studies collected saliva specimens in sterile containers or cups whereas one study used wide plastic tubes. Reverse transcription polymerase chain reaction (RT-PCR) assays were used in all studies, with eight studies utilising commercial assays and two studies utilising in-house laboratory-developed and validated assays. An overview of all study characteristics is summarised in **Table 1**; characteristics related to sample collection and testing are summarised in **Table 2**. The QUADAS-2 tabular summary and graphical summary is provided in the supplementary material *(p 9)*. Additionally, a short summary of each the studies included and the 2×2 contingency tables are provided in supplementary *(p 4)*.

**Table 2.**
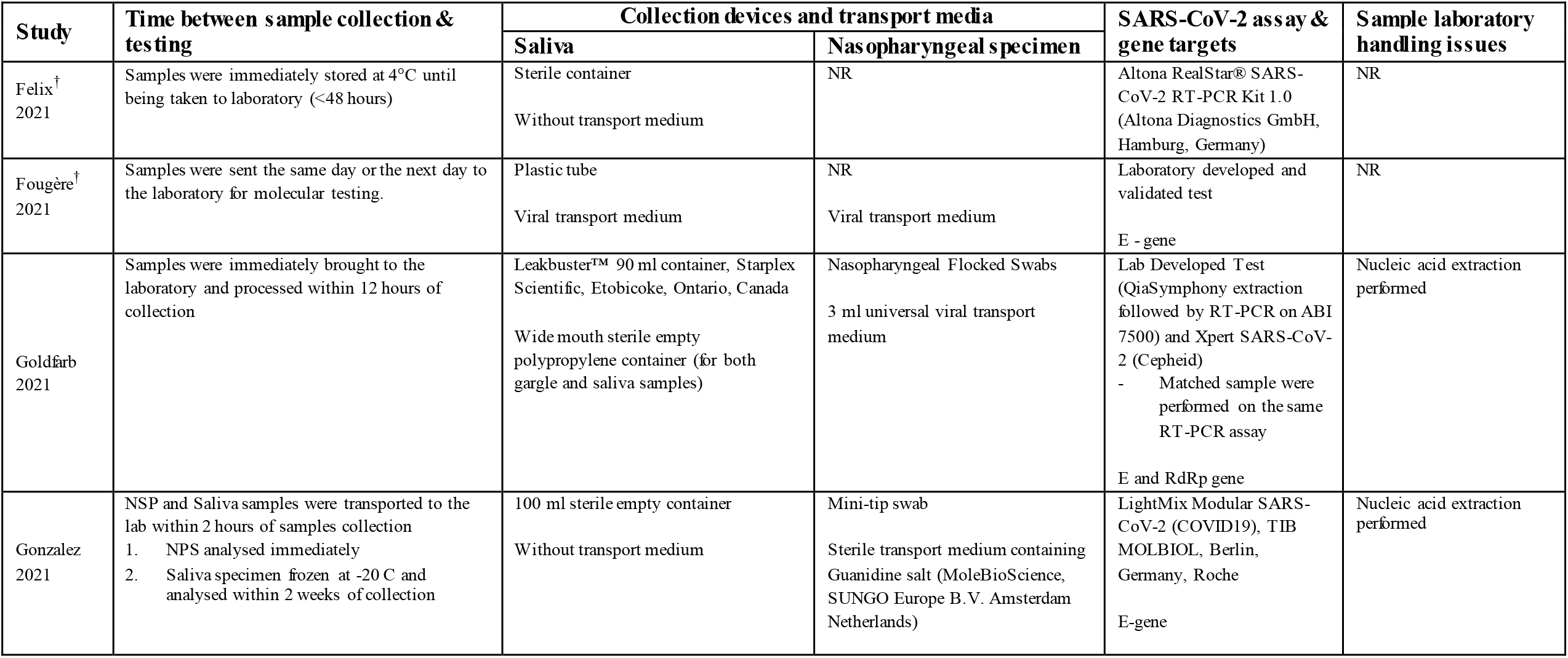
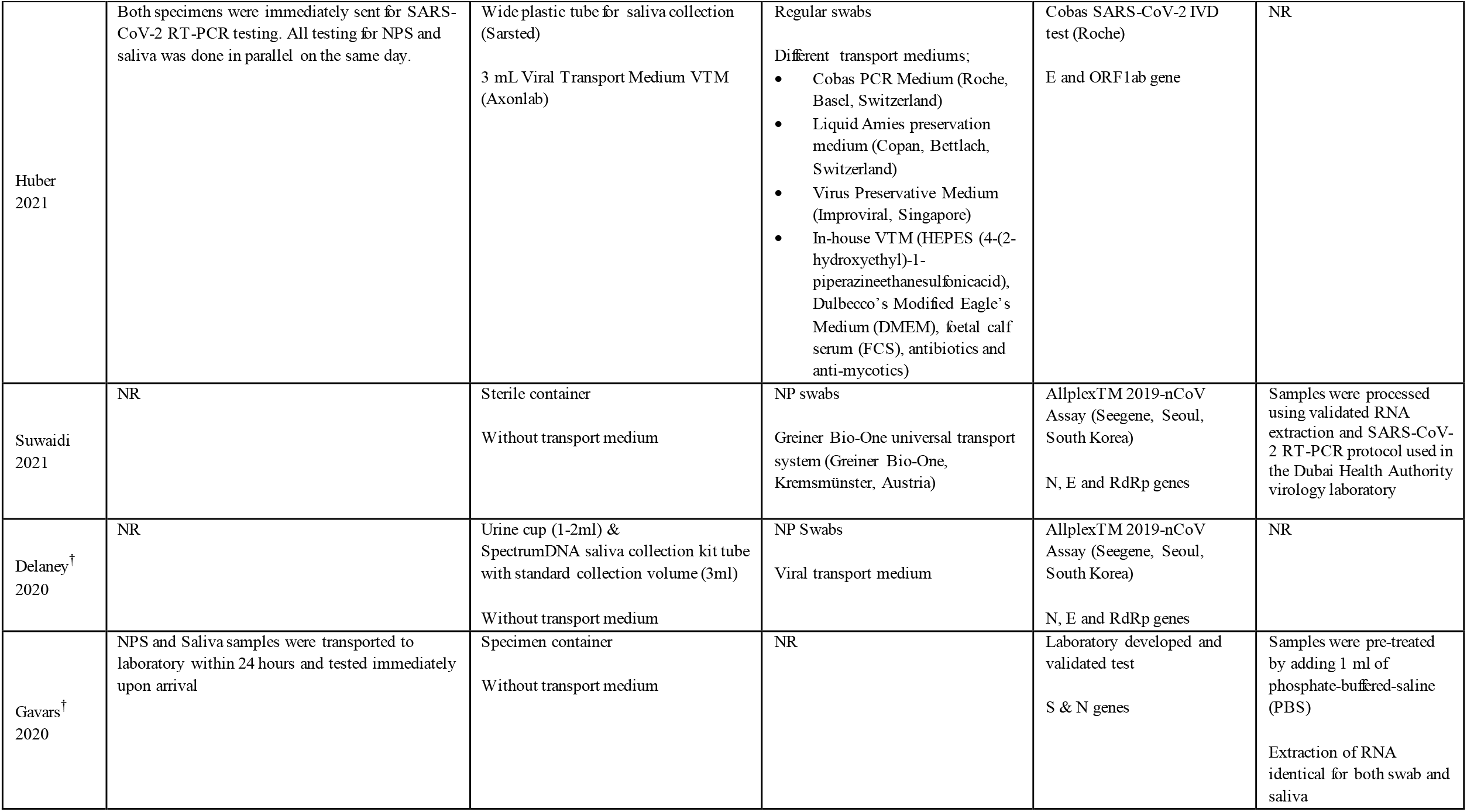
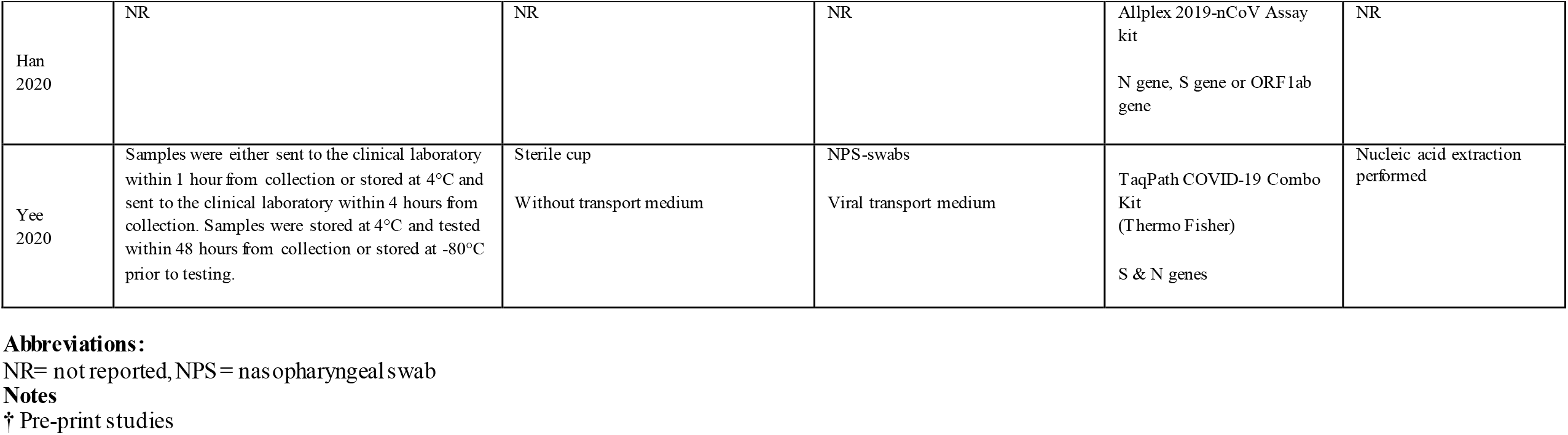
Characteristics related to collection and testing of included studies

### Meta-analysis

In total, we included 1486 paired saliva and nasopharyngeal samples obtained from children ages 1-18 years old. In the meta-analysis, we obtained a pooled absolute saliva sensitivity of 84.5% (95% CI; 78.0%-90.3%) Heterogeneity I^2^ was 23%. The pooled absolute specificity was 99.5% (95% CI; 98.2%-100.0%). Individual study estimates of sensitivity and specificity can be seen in Figure 2. The highest sensitivity of 93.3% (95% CI; 78.7%-98.2%) was observed by Huber et al where saliva was collected by asking asymptomatic and symptomatic children to clear their throat to include sample material from the back of the oropharynx for specimen collection. Stratified comparison of symptom status did not show a significant difference in sensitivity. Table 3 shows results stratified by different study characteristics. Often stratifying information was not reported. None of the stratifying variables had a significant impact on the accuracy estimates.

**Figure 2.**
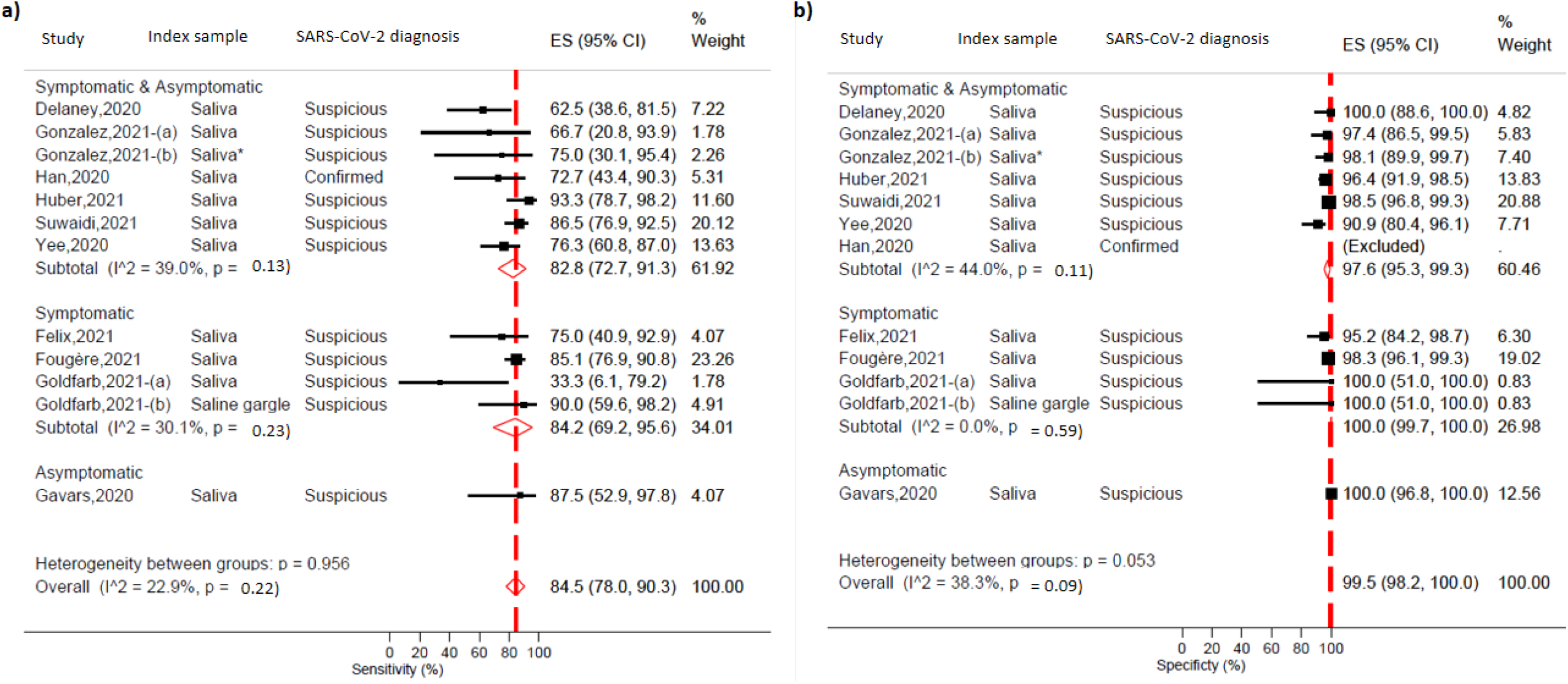
**a)** Absolute sensitivity and **b)** absolute specificity of saliva in children (<18 years old) using RT-PCR on nasopharyngeal samples as reference standard stratified by symptomatic status Note: * Saliva samples were collected after oropharyngeal washing ** Han et all was excluded in the specificity analysis as there were no negative cases

**Table 3.** Stratified pooled estimates of saliva for SARS-CoV-2 detection using RT-PCR on nasopharyngeal samples as a reference in children by study characteristics

| Study Characteristic | Studies, n | Sensitivity (95% CI) | Specificity (95% CI) |
| --- | --- | --- | --- |
| **Population** |  |  |  |
| Symptomatic | 3 | 84.2 (52.9-97.8) | 100.0 (99.7-100.0) |
| Asymptomatic | 1 | 87.5 (52.9-97.8) | 100.0 (96.8-100.0) |
| Symptomatic and asymptomatic | 6 | 82.8 (72.7-91.3) | 97.6 (95.3-99.3) |
| Heterogeneity between groups |  | p=1.0 | p=0.1 |
| **SARS-CoV-2 diagnosis** |  |  |  |
| Suspicious | 9 | 85.1 (78.3-91.0) | 99.5 (98.2-100.0) |
| Confirmed | 1 | 72.7 (43.4-90.3) | ** |
| Heterogeneity between groups |  | p=0.4 | - |
| **Peer-reviewed** |  |  |  |
| Yes | 6 | 85.6 (76.3-93.2) | 99.3 (97.4-100.0) |
| No | 4 | 80.8 (68.0-91.4) | 99.3 (97.1-100.0) |
| Heterogeneity between groups |  | p=0.7 | p=0.2 |
| **Index sample\*** |  |  |  |
| Saliva | 10 | 83.6 (76.2-90.2) | 99.1 (76.2-100.0) |
| Oropharyngeal washing | 1 | 75.0 (30.1-95.4) | 100.0 (51.0-100.0) |
| Saline gargle | 1 | 90.0 (59.6-98.2) | 98.1(89.9-99.7) |
| Heterogeneity between groups |  | p=0.8 | p=0.9 |
| **Reference sample** |  |  |  |
| Nasopharyngeal samples | 8 | 86.5 (80.5-91.8) | 99.4 (98.0-100.0) |
| Naso/oropharyngeal sample | 2 | 71.6 (50.9-89.0) | 100.0 (99.0-100.0) |
| Heterogeneity between groups |  | p=0.2 | p=0.01 |
| **Saliva transport medium** |  |  |  |
| Unpreserved | 4 | 88.4 (78.9-95.9) | 100.0 (99.9-100.0) |
| Viral Transport Medium | 2 | 87.4 (81.0-92.8) | 97.8 (96.2-99.0) |
| Unreported | 4 | 73.2 (61.8-83.4) | 95.9 (88.9-99.8) |
| Heterogeneity between groups |  | p=0.03 | p=0.2 |
| *2 studies included 2 datasets each with different saliva sampling technique |  |  |  |
| ** No specificity estimate since the study was restricted to confirmed SARS-CoV-2 carriers (as defined by presence of viral RNA in nasopharyngeal specimens) |  |  |  |

We also performed a post hoc analysis restricted to peer-reviewed studies *(Supplementary Figure 2, p11)* and found a slightly higher pooled sensitivity of 88.5% (95% CI; 80.9%-94.8%) and a similar specificity of 99.3 (95% CI; 97.7%-100.0%).

## Discussion

To our knowledge, this is the first systematic review and meta-analysis on the accuracy of SARS-CoV-2 assays applied on paired saliva and nasopharyngeal swabs in children. We addressed a highly relevant public health concern when it comes to testing for SARS-CoV-2 in children. We identified 10 studies with primary data extracted from original articles as well as data provided directly by authors. In our meta-analysis, we found a pooled sensitivity of 84.5% and a pooled specificity of 99.5% for SARS-CoV-2 detection in saliva with RT-PCR using nasopharyngeal samples as reference. We were unable to demonstrate that the performance of saliva varied by symptom status. However, given the lack of symptom information, we cannot ascertain that symptom status does influence the sensitivity of SARS-CoV-2 testing on saliva. We, therefore, plan on contacting authors to provide more information about symptomatic status.

The results from our meta-analysis show that the sensitivity and specificity of saliva sampling in children are similar to adults and the general population. Previous systematic reviews and meta-analysis in adults showed pooled sensitivity ranges of 83.2%-86.9% and specificity of 98.9%^22,23^. This shows that the accuracy of saliva for the detection of SARS-CoV-2 in children is similar to adults.

Limitations of our study are the lack of data stratified by symptoms and stage of infection in the included studies. It has been demonstrated that the stage of infection during sample collection can contribute to test sensitivity differences. It would have been valuable to compare the sensitivity of saliva in more detail in symptomatic and asymptomatic children. Many studies in our meta-analysis included both symptomatic and asymptomatic children in their studies without clear stratification. Besides, most of the studies included an outpatient population. Studies that included critically ill and hospitalised children were lacking and hence our findings may not be applicable to this setting. Additionally, we assumed nasopharyngeal samples as the reference standard which is an imperfect diagnostic test. Some studies have shown that SARS-CoV-2 remained detectable in saliva while NPS tested negative^24,25^.

In conclusion, we found that the accuracy of saliva for the detection of SARS-CoV-2 by RT-PCR in children was similar to results seen in the adult population.

## Data Availability

Not applicable

## Acknowledgements

SKD and MA would like to acknowledge the VALCOR^26^ project funded by emergency funding from the Federal Belgian Government.

We thank the following authors for providing us with additional data and information that lead to the success of this meta-analysis;

Dr Didzis Gavars (*E. Gulbja Laboratory, Riga, Latvia*),

Dr David Goldfarb (*Associate Head at the Department of Pathology & Lab Medicine, BC Children’s & Women’s Hospital*),

Dr Félix Gutiérrez Rodero (*Professor of Medicine at the Infectious Diseases Unit Hospital General Universities d’
sElx & Universidad Miguel Hernández Alicante, Spain*),

Dr Meghan Delaney (*Director, Transfusion Medicine and Division Chief, Pathology & Laboratory Medicine Division from the Children’s National Hospital, Washington USA*),

Dr Joseph Campos (*Director of Microbiology Laboratory from the Children’s National Hospital, Washington USA*) and

Dr Jennifer Dien Bard

(*Director, Clinical Microbiology and Virology Department of Pathology and Laboratory Medicine, Children’s Hospital Los Angeles, USA*).

