## Supplementary material for "The accuracy of saliva versus nasopharyngeal and/or oropharyngeal samples for the detection of SARS-CoV-2 in children – A rapid systematic review and meta-analysis"

#### Clinical questions and PICOTS components study question

**Study question:**
What is the diagnostic accuracy of saliva as compared to the standard naso- and /or oro-pharyngeal samples for the detection of SARS-CoV-2 in children?

**PICOTS components**
***Population***: Children (age <18 years old) being tested for SARS-CoV-2 for current SARS-CoV-2 infection or children undergoing screening for SARS-CoV-2 infection (asymptomatic and/or screening)
***Intervention test***: Saliva samples tested on the same RT-PCR for naso- and/or oro-pharyngeal swabs
***Comparator tests***: Naso- and/or oro-pharyngeal swabs tested on the same RT-PCR for saliva samples
***Outcomes***:
O1: absolute sensitivity and specificity of saliva compared to naso- and/or oro-pharyngeal swabs for the detection of SARS-CoV-2
***Target condition:***
Detection of SARS-CoV-2 infection
***Studies***:
Cross-sectional, cohort, randomized clinical trials, case-controls diagnostic test accuracy studies where a matched samples (saliva and naso- and/or oro-pharyngeal swabs) were taken from the same individual and tested on the same RT-PCR platform.

#### Literature retrieval strings

**A. In Pubmed-Medline**

(saliva) and (corona*[Title] or covid*[Title] or sars*[Title] or "2019-nCoV"[Title]) and (accuracy or sensitiv* or specific* or "detection limit" or yield or positiv* or copies or "copy number*" or lod or "viral load*" or "viral titre*" or "viral culture*" or "viral rna" or "limit of detection" or "predictive value" or "cycle threshold" or ppv or npv)

**B. Live Evidence**

Saliva and (child* or pead* or pedi*) and (accuracy or sensitiv* or specific)

#### Brief Study Descriptions and 2x2 Contingency Tables

The contingency tables report the comparison of saliva with nasopharyngeal samples (or oropharyngeal samples).

**Felix[1]**

This study evaluated saliva samples from 50 children with suspected COVID-19 who attended public healthcare services. Children were asked to spit into a sterile container for the collection of about 1ml of saliva after the NPS collection. Children were described to have mild symptoms. Information on viral load and threshold cycle was unavailable.

This study is considered to be an uncertain risk of bias as we could not determine the patient flow and if patient selection criteria introduced bias. There was not much information in the article.

| | | **NPS** | | |
|---|---|---|---|---|
| | | Positive | Negative | *Total* |
| **Saliva** | Positive | 6 | 2 | 8 |
| | Negative | 2 | 40 | 42 |
| | *Total* | 8 | 42 | 50 |

**Fougère[2]**

This was an observational prospective multicentre comparative study where children and adolescents ages 1 month to 18 years were recruited from two separate outpatient clinics. Patients were all symptomatic. Saliva samples were collected either by a healthcare worker, the patient itself or under caregiver supervision. Saliva and NP samples collected and sent the same day or the next morning to the molecular diagnostics laboratory for RT-PCR analyses. This study excluded asymptomatic and included only outpatients and not hospitalized patients.

The likelihood of patient selection bias was high as there was the inclusion of subjects who may have been known to be positive for COVID-19.

| | | **NPS** | | |
|---|---|---|---|---|
| | | Positive | Negative | *Total* |
| **Saliva** | Positive | 86 | 5 | 91 |
| | Negative | 15 | 291 | 306 |
| | *Total* | 101 | 296 | 397 |

**Goldfarb[3]**

In this study, symptomatic outpatient children presenting at a testing centre in a hospital(4-12 years old) were enrolled. Saline gargle and/or saliva samples were collected in addition to a NPS.

For children who provided both saliva and saline mouth rinse/gargle samples the order of sample collection was alternated sequentially (i.e. saliva first vs mouth rinse/gargle sample first). Saliva samples were self-collected (or aided by parent/caregiver). All samples were collected at the same time. Participants who had all 3 samples collected were asked about the acceptability of each sample type. Saliva samples that were deemed to have too much mucous by the processing technologist were diluted 1:1 in TE buffer. Data was requested from the author for children strata however it was not clear how many children provided an only saliva sample and/or gargle samples.

| | | NPS | | |
|---|---|---|---|---|
| | | Positive | Negative | *Total* |
| **Saliva** | Positive | 1 | 0 | 1 |
| | Negative | 2 | 4 | 6 |
| | *Total* | 3 | 4 | 7 |

| | | NPS | | |
|---|---|---|---|---|
| | | Positive | Negative | *Total* |
| **Gargle samples** | Positive | 9 | 0 | 9 |
| | Negative | 1 | 4 | 5 |
| | *Total* | 10 | 4 | 14 |

**Gonzalez[4]**

This paper reports the prospective collection of three different saliva specimens in an ambulatory setting (outpatients with a broad clinical spectrum of disease, including asymptomatic cases) undergoing SARS-CoV-2 testing. Patients were randomized to either one of the saliva collection methods. Different collection methods were a) saliva under supervised collection (where a healthcare worker was present to supervise saliva collection) b) saliva specimens after oropharyngeal washing c) self-collected saliva. A matched NPS sample was collected after saliva collection. The author was contacted to provide data for the children cohort and data was received for saliva and saliva oropharyngeal washing. Paired samples were tested on the same PCR platform. Ct value of ≤40 by PCR is considered a positive result. 44 children were unable to provide saliva specimens (mean age 5.8 years). In all the patients (adults & children), the supervised collection method had higher sensitivity compared to the self-collection and saliva after oropharyngeal washing.

The likelihood of bias was low due to recruitment. The likelihood of applicability bias was high as the study included small sample size and the difference in performance among collection methods may be confounded by differences in population. Besides a large number of children were unable to provide saliva samples for testing.

| | | NPS | | |
|---|---|---|---|---|
| | | Positive | Negative | *Total* |
| **Oropharyngeal washing** | Positive | 3 | 1 | 4 |
| | Negative | 1 | 51 | 52 |
| | *Total* | 4 | 52 | 56 |

| | | NPS | | |
|---|---|---|---|---|
| | | Positive | Negative | *Total* |
| | Positive | 2 | 1 | 3 |
| **Saliva** | Negative | 1 | 37 | 38 |
| | *Total* | 3 | 38 | 41 |

**Huber[5]**

This study included adults and children opting for SARS-CoV-2 testing at five test sites. Four test sites were exclusive for outpatients (three for adults, one for children) and one test was in an emergency department. Individuals presenting at test centres were either symptomatic or had been recently exposed to a positive COVID-19 case. Saliva was collected by asking patients to clear their throat to include sampling material from the posterior oropharynx. Saliva was self-collected. Matched NPS and saliva samples were collected and both samples were identically processed on the same day in the laboratory. Data was available online.

This study is believed to have a low risk of bias.

| | | NPS | | |
|---|---|---|---|---|
| | | Positive | Negative | *Total* |
| **Saliva** | Positive | 28 | 5 | 33 |
| | Negative | 2 | 135 | 137 |
| | *Total* | 30 | 140 | 140 |

**Suwaidi[6]**

This was a prospective observational diagnostic study specific to school children. It included a population-based convenience sample of school children attending COVID-19 screening at a health authority screening centre. Children were being tested if they had been in contact with a confirmed COVID-19 patient, presenting with symptoms and/or children returning to school. Matched NPS and saliva were collected at the same time. Saliva was self-collected and 1-3ml was obtained. A cycle threshold of ≤40 was taken as the cut-off for a positive result for the target genes as per manufacturer-provided protocol. A total of 476 children with a mean age of 10.8 years were included. There were 485 pairs of NPS and saliva matched samples (9 children had repeated sampling and provided two sets of paired samples due to clinical reassessment). Results were provided for both matched 476 samples and 485 matched samples.

This study is believed to have a low risk of bias.

| | | NPS | | |
|---|---|---|---|---|
| | | Positive | Negative | *Total* |
| **Saliva** | Positive | 64 | 6 | 70 |
| | Negative | 10 | 396 | 406 |
| | *Total* | 74 | 402 | 476 |

**Delaney**[7]

A prospective study of symptomatic and asymptomatic paediatric patients from an urban paediatric, tertiary medical centre were tested for SARS-CoV-2 and provided nasopharyngeal or oropharyngeal samples. A matched saliva sample was also obtained using two different collection device; a urine cup and a commercially EUA- approved SpectrumDNA saliva collection to determine which collection device was more favourable for sample collection and testing. Although the article did not include data on the diagnostic accuracy of the RT-PCR tests, the author provided us with data when requested. All matched samples were tested on the same RT-PCR platform. Stratified data on symptom status or saliva collection device was unavailable.

This study is considered to be an uncertain risk of bias as we could not determine the patient flow and if patient selection criteria introduced bias. There was not much information in the article.

| | | **Naso-/oro-pharyngeal sample** | | |
|---|---|---|---|---|
| | | Positive | Negative | *Total* |
| **Saliva** | Positive | 10 | 0 | 10 |
| | Negative | 6 | 30 | 36 |
| | *Total* | 16 | 30 | 46 |

**Gavars**[8]

This study was a single-centre study that included adults and children attending a SARS-CoV-2 testing centre who were either symptomatic or had been in close contacts with a confirmed COVID-19 patient. Saliva samples were self-collected and patients were requested to collect saliva in an isolated location (outdoors or in their car). Samples were tested on the same RT-PCR platform. The author was contacted to provide data specifically on children strata.

The likelihood of patient selection bias was considered to be low.

| | | **NPS** | | |
|---|---|---|---|---|
| | | Positive | Negative | *Total* |
| | Positive | 7 | 0 | 7 |
| **Saliva** | Negative | 1 | 117 | 118 |
| | *Total* | 8 | 117 | 125 |

**Han**[9]

This study included confirmed COVID-19 positive children (<18 years) who were hospitalised. Children enrolled in this study were either symptomatic or asymptomatic. There were 12 patients enrolled however saliva sample were only collected from 11 patients. Details on why one patient did not provide saliva sample were not reported. Saliva collection method, device and transport media used was not reported. SARS-CoV-2 was detected in saliva in the early phases of disease onset (0-7 days after symptoms appear).

The likelihood of patient selection bias was high as there was inclusion of subjects who may have been known to be positive for COVID-19.

| | | NPS | | |
|---|---|---|---|---|
| | | Positive | Negative | *Total* |
| **Saliva** | Positive | 8 | 0 | 8 |
| | Negative | 3 | 0 | 3 |
| | *Total* | 11 | 0 | 11 |

**Yee[10]**

This study enrolled children and adults from a single medical institution that included outpatients and patients admitted to the emergency department. Patients who had previously tested positive for COVID-19 as well individuals with unknown COVID-19 status were included. SARS-CoV-2. Both symptomatic and asymptomatic patients were enrolled in the study. Saliva was self-collected under the supervision of a healthcare worker and NPS sample was subsequently collected. A positive result was determined by using a cut-off threshold cycle (CT) value of ≤40. The author was contacted to provide data specifically on children strata.

The likelihood of patient selection bias was high as there was the inclusion of subjects who may have been known to be positive for COVID-19.

| | | NPS | | |
|---|---|---|---|---|
| | | Positive | Negative | *Total* |
| **Saliva** | Positive | 29 | 5 | 34 |
| | Negative | 9 | 50 | 59 |
| | *Total* | 38 | 55 | 93 |

#### Quality Assessment of Diagnostic Accuracy Studies (QUADAS)

**Table S1**. Risk of bias in included studies

| | Risk of bias items | | | | Applicability with regard to review question and context | | |
|---|---|---|---|---|---|---|---|
| **Study** | **Patient selection** | **Index test** | **Reference standard** | **Flow and timing** | **Patient Selection** | **Index test** | **Reference standard** |
| Gonzalez 2021 | Low | Unclear | Low | Low | High | Low | Low |
| Gavars 2020 | Low | Low | Low | Low | Low | Unclear | Low |
| Yee 2020 | High | Low | Low | Low | High | Low | Low |
| Han 2020 | High | High | Low | Low | High | Unclear | Unclear |
| Goldfarb 2021 | Low | Low | Low | Low | Low | Unclear | Low |
| Suwaidi 2021 | Low | Unclear | Low | Low | Low | Low | Low |
| Delaney 2021 | Unclear | Unclear | Unclear | High | Unclear | Unclear | Unclear |
| Huber 2021 | Unclear | Low | Low | Unclear | Low | Unclear | Low |
| Felix 2021 | High | Unclear | Low | Low | Unclear | Low | Low |
| Fougère 2021 | High | Unclear | Low | High | High | Low | Low |

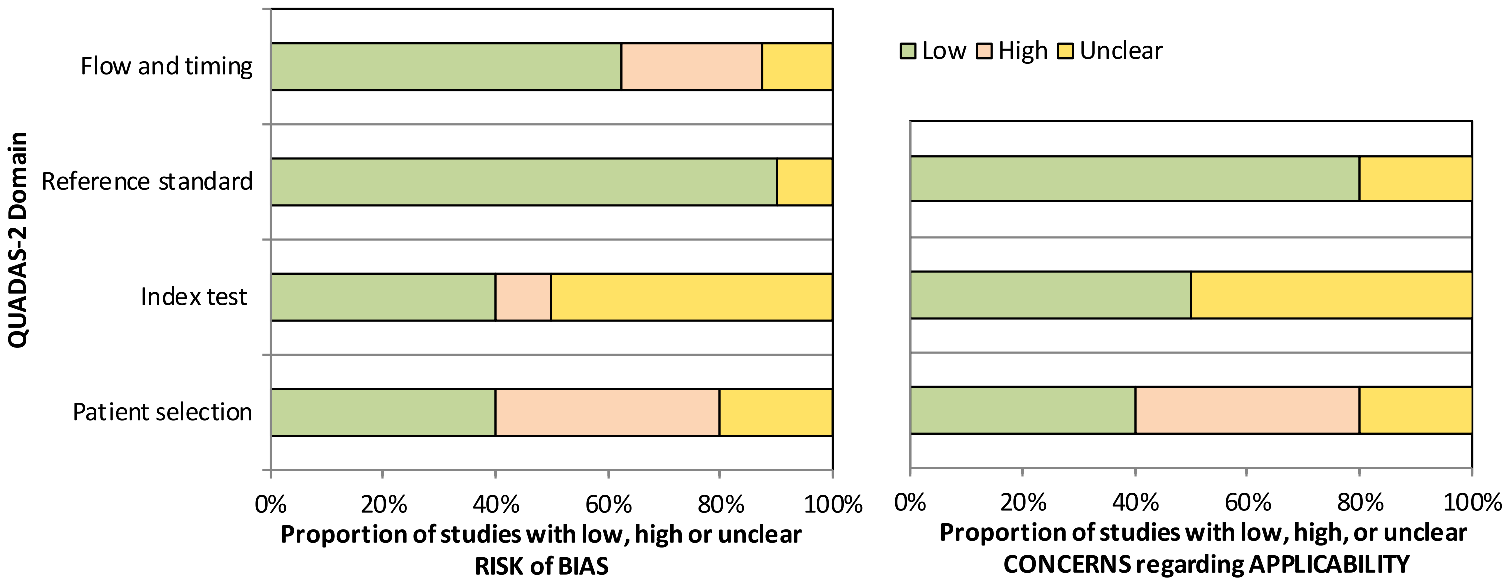

**Figure S1**. Quality Assessment of Diagnostic Accuracy Studies 2 (QUADAS-2) Study Quality Summary

### Sub-group meta-analyses

a)

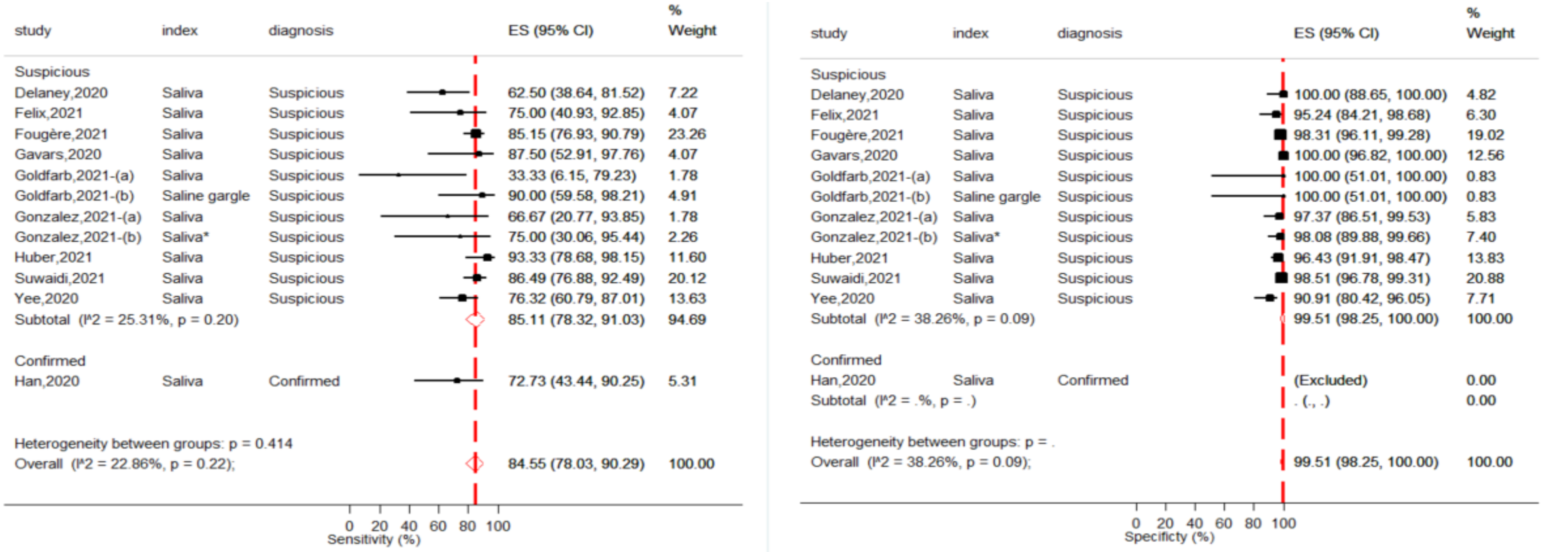

b)

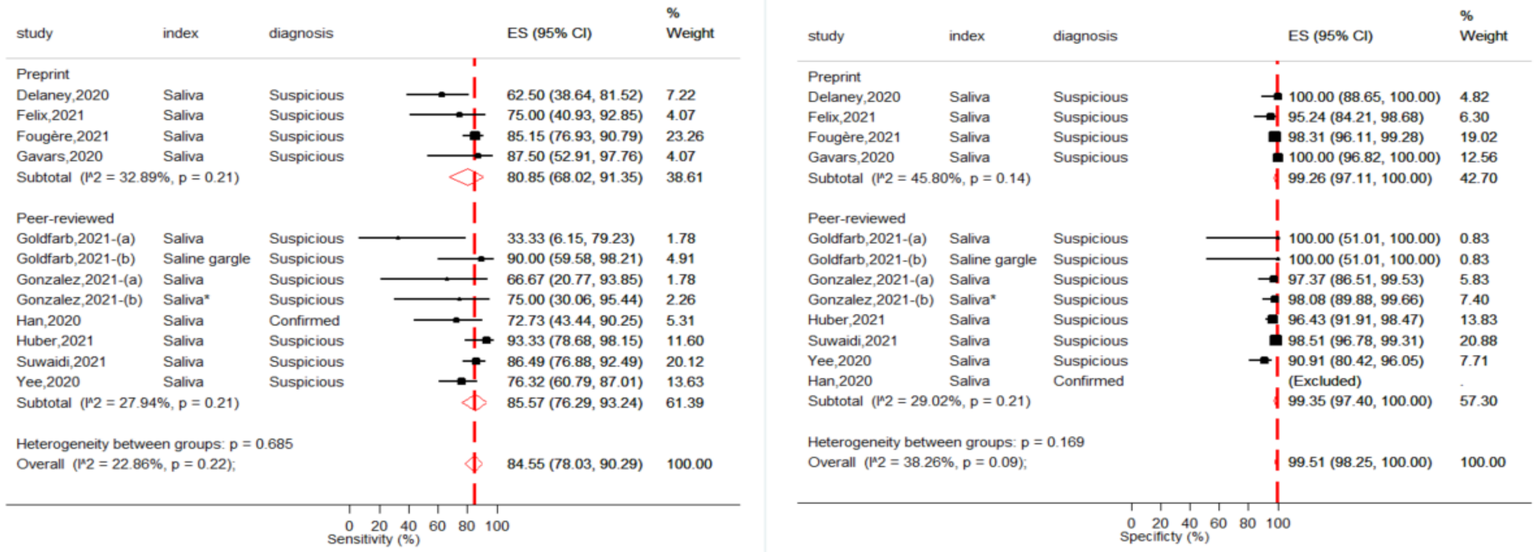

c)

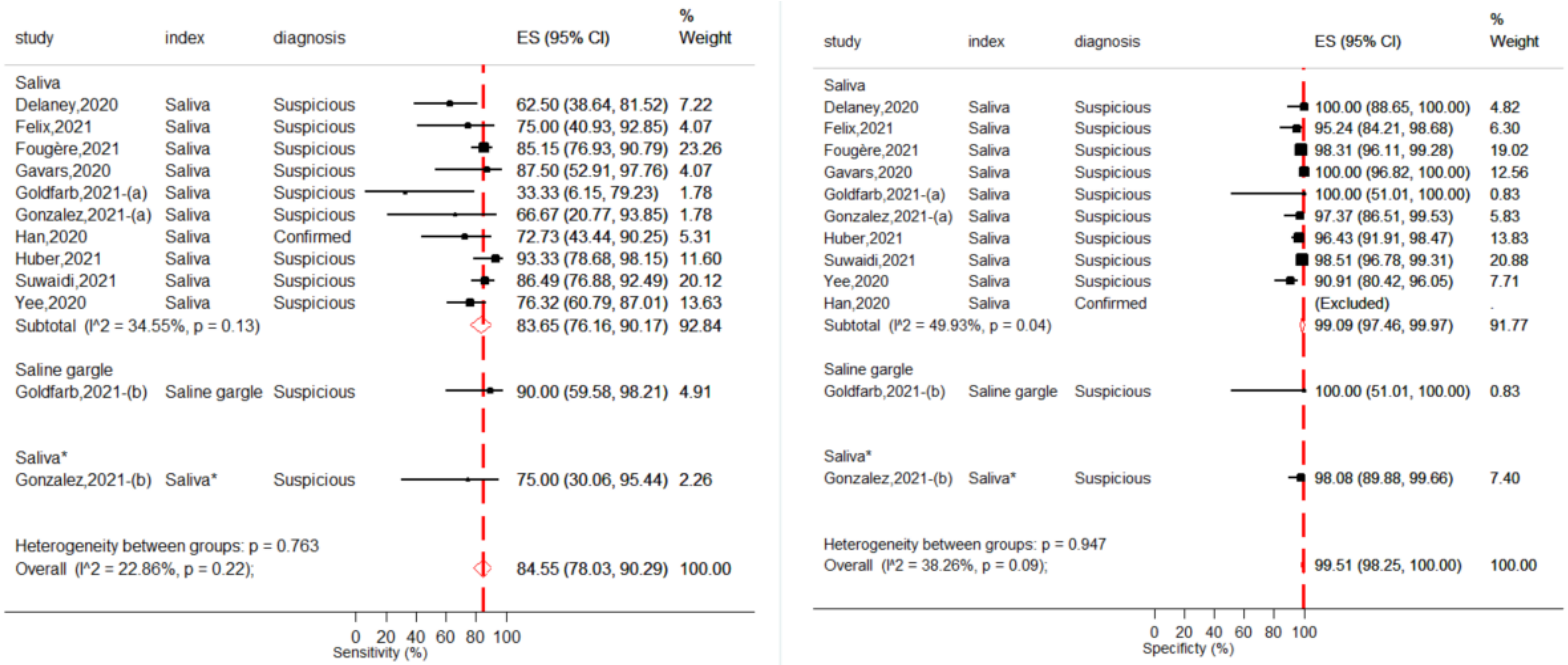
study index diagnosis ES (95% CI) % Weight
Saliva
Delaney,2020 Saliva Suspicious 62.50 (38.64, 81.52) 7.22
Felix,2021 Saliva Suspicious 75.00 (40.93, 92.85) 4.07
Fougère,2021 Saliva Suspicious 85.15 (76.93, 90.79) 23.26
Gavars,2020 Saliva Suspicious 87.50 (52.91, 97.76) 4.07
Goldfarb,2021-(a) Saliva Suspicious 33.33 (6.15, 79.23) 1.78
Gonzalez,2021-(a) Saliva Suspicious 66.67 (20.77, 93.85) 1.78
Han,2020 Saliva Confirmed 72.73 (43.44, 90.25) 5.31
Huber,2021 Saliva Suspicious 93.33 (78.68, 98.15) 11.60
Suwaidi,2021 Saliva Suspicious 86.49 (76.88, 92.49) 20.12
Yee,2020 Saliva Suspicious 76.32 (60.79, 87.01) 13.63
Subtotal (I^2 = 34.55%, p = 0.13) 83.65 (76.16, 90.17) 92.84
Saline gargle
Goldfarb,2021-(b) Saline gargle Suspicious 90.00 (59.58, 98.21) 4.91
Saliva*
Gonzalez,2021-(b) Saliva* Suspicious 75.00 (30.06, 95.44) 2.26
Heterogeneity between groups: p = 0.763
Overall (I^2 = 22.86%, p = 0.22); 84.55 (78.03, 90.29) 100.00
0 20 40 60 80 100
Sensitivity (%)
study index diagnosis ES (95% CI) % Weight
Saliva
Delaney,2020 Saliva Suspicious 100.00 (88.65, 100.00) 4.82
Felix,2021 Saliva Suspicious 95.24 (84.21, 98.68) 6.30
Fougère,2021 Saliva Suspicious 98.31 (96.11, 99.28) 19.02
Gavars,2020 Saliva Suspicious 100.00 (96.82, 100.00) 12.56
Goldfarb,2021-(a) Saliva Suspicious 100.00 (51.01, 100.00) 0.83
Gonzalez,2021-(a) Saliva Suspicious 97.37 (86.51, 99.53) 5.83
Huber,2021 Saliva Suspicious 96.43 (91.91, 98.47) 13.83
Suwaidi,2021 Saliva Suspicious 98.51 (96.78, 99.31) 20.88
Yee,2020 Saliva Suspicious 90.91 (80.42, 96.05) 7.71
Han,2020 Saliva Confirmed (Excluded) .
Subtotal (I^2 = 49.93%, p = 0.04) 99.09 (97.46, 99.97) 91.77
Saline gargle
Goldfarb,2021-(b) Saline gargle Suspicious 100.00 (51.01, 100.00) 0.83
Saliva*
Gonzalez,2021-(b) Saliva* Suspicious 98.08 (89.88, 99.66) 7.40
Heterogeneity between groups: p = 0.947
Overall (I^2 = 38.26%, p = 0.09); 99.51 (98.25, 100.00) 100.00
0 20 40 60 80 100
Specificty (%)

d)

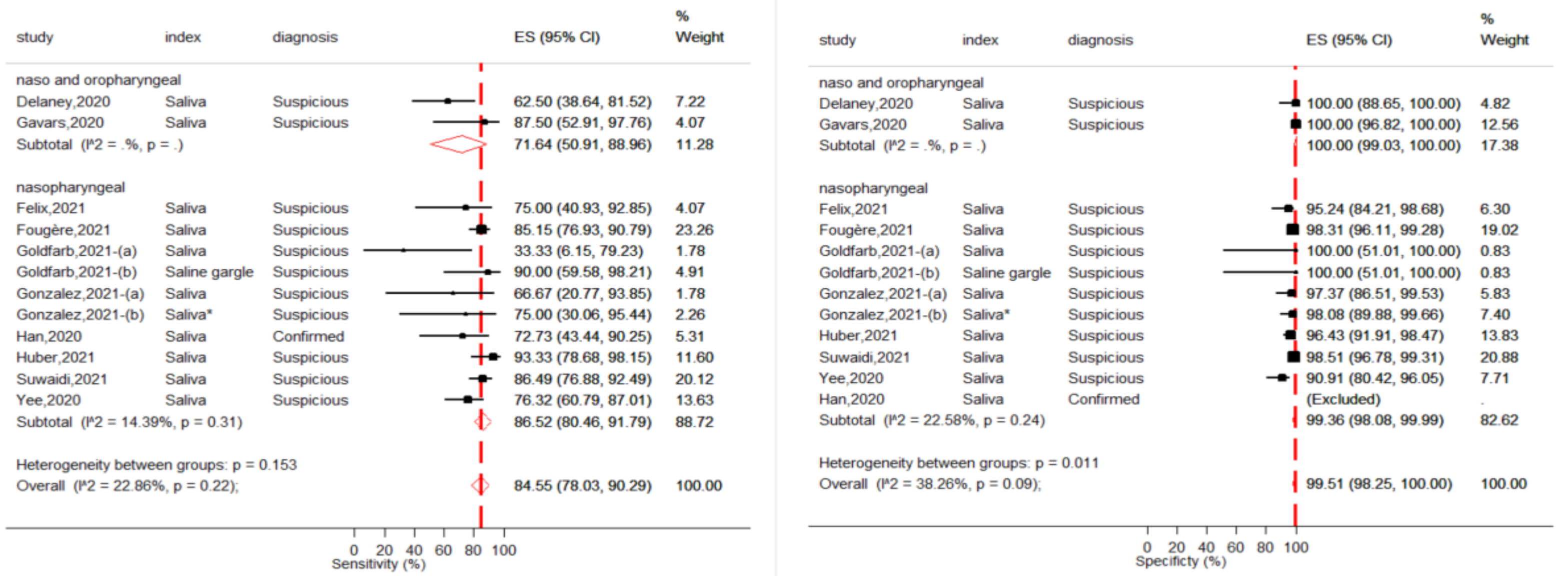
study index diagnosis ES (95% CI) % Weight
naso and oropharyngeal
Delaney,2020 Saliva Suspicious 62.50 (38.64, 81.52) 7.22
Gavars,2020 Saliva Suspicious 87.50 (52.91, 97.76) 4.07
Subtotal (I^2 = .%, p = .) 71.64 (50.91, 88.96) 11.28
nasopharyngeal
Felix,2021 Saliva Suspicious 75.00 (40.93, 92.85) 4.07
Fougère,2021 Saliva Suspicious 85.15 (76.93, 90.79) 23.26
Goldfarb,2021-(a) Saliva Suspicious 33.33 (6.15, 79.23) 1.78
Goldfarb,2021-(b) Saline gargle Suspicious 90.00 (59.58, 98.21) 4.91
Gonzalez,2021-(a) Saliva Suspicious 66.67 (20.77, 93.85) 1.78
Gonzalez,2021-(b) Saliva* Suspicious 75.00 (30.06, 95.44) 2.26
Han,2020 Saliva Confirmed 72.73 (43.44, 90.25) 5.31
Huber,2021 Saliva Suspicious 93.33 (78.68, 98.15) 11.60
Suwaidi,2021 Saliva Suspicious 86.49 (76.88, 92.49) 20.12
Yee,2020 Saliva Suspicious 76.32 (60.79, 87.01) 13.63
Subtotal (I^2 = 14.39%, p = 0.31) 86.52 (80.46, 91.79) 88.72
Heterogeneity between groups: p = 0.153
Overall (I^2 = 22.86%, p = 0.22); 84.55 (78.03, 90.29) 100.00
0 20 40 60 80 100
Sensitivity (%)
study index diagnosis ES (95% CI) % Weight
naso and oropharyngeal
Delaney,2020 Saliva Suspicious 100.00 (88.65, 100.00) 4.82
Gavars,2020 Saliva Suspicious 100.00 (96.82, 100.00) 12.56
Subtotal (I^2 = .%, p = .) 100.00 (99.03, 100.00) 17.38
nasopharyngeal
Felix,2021 Saliva Suspicious 95.24 (84.21, 98.68) 6.30
Fougère,2021 Saliva Suspicious 98.31 (96.11, 99.28) 19.02
Goldfarb,2021-(a) Saliva Suspicious 100.00 (51.01, 100.00) 0.83
Goldfarb,2021-(b) Saline gargle Suspicious 100.00 (51.01, 100.00) 0.83
Gonzalez,2021-(a) Saliva Suspicious 97.37 (86.51, 99.53) 5.83
Gonzalez,2021-(b) Saliva* Suspicious 98.08 (89.88, 99.66) 7.40
Huber,2021 Saliva Suspicious 96.43 (91.91, 98.47) 13.83
Suwaidi,2021 Saliva Suspicious 98.51 (96.78, 99.31) 20.88
Yee,2020 Saliva Suspicious 90.91 (80.42, 96.05) 7.71
Han,2020 Saliva Confirmed (Excluded) .
Subtotal (I^2 = 22.58%, p = 0.24) 99.36 (98.08, 99.99) 82.62
Heterogeneity between groups: p = 0.011
Overall (I^2 = 38.26%, p = 0.09); 99.51 (98.25, 100.00) 100.00
0 20 40 60 80 100
Specificty (%)

e)

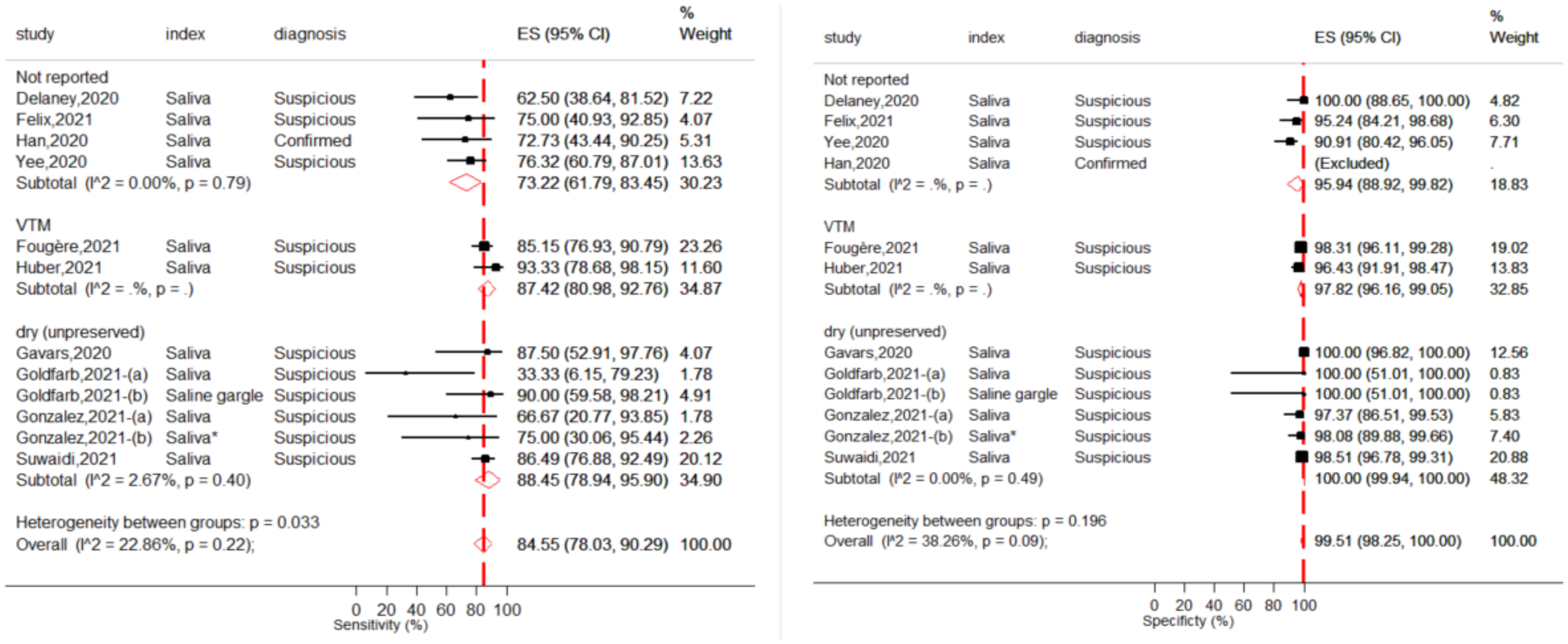

**Figure S2**. Absolute sensitivity and absolute specificity of SARS-CoV-2 RT PCR on saliva in children (<18 years old) using RT-PCR on nasopharyngeal samples as reference standard by **a)** SARS-CoV-2 diagnosis **b)** Peer-reviewed and preprints **c)** Index samples **d)** Reference sample **e)** Saliva transport medium

Note: * Saliva samples were collected after oropharyngeal washing
